# Development of a novel medical physics patient consult program

**DOI:** 10.1101/2020.06.24.20135061

**Authors:** Bradley W Schuller, Jonathan A Baldwin, Elizabeth A Ceilley, Alexander Markovic, Jeffrey M Albert

**Affiliations:** Banner MD Anderson Cancer Center at McKee Medical Center, Loveland, CO USA; Banner MD Anderson Cancer Center at North Colorado Medical Center, Greeley, CO USA

**Author notes:** Corresponding Author: Bradley W Schuller PhD.

**Keywords:** patient education, patient satisfaction, health literacy

## Abstract

**Purpose:** To develop a new patient consult program, where patients are invited to meet directly with a clinical medical physicist to learn and ask questions about the technical aspects of their care.

**Methods:** Patients are invited to meet voluntarily with a clinical medical physicist directly after the treatment planning CT appointment, and then again after treatment starts. Each consult starts with an overview of the clinical medical physicist’s role in patient care. This is followed by a detailed explanation of the treatment planning CT, treatment planning, and treatment delivery processes. Data are collected after each patient encounter, including: age, gender, treatment intent, treatment site, consult duration, discussion points, overall impression, and a summary of the questions asked. Qualitative data analysis focused on understanding the number and types of questions asked during the physics consults. Additional analyses focused on evaluating the encounter notes for interesting insights regarding meeting tone, number of meeting attendees, and other non-clinical discussion points.

**Results:** Sixty three patients were seen between August 2016 and December 2017, accounting for 29% of the total department patient load. The average physics consult duration was 24 minutes. When evaluating the patient encounter notes for overall tone, 55 patients (87%) had positive descriptors such as “pleasant conversation”. Thirty three patients (52%) brought at least one other person into the consult, and 27 patients (43%) contributed personal stories or professional background information to the conversation. When the collection of patient questions was grouped into question types, the data show that the majority of the consult discussion addresses questions related to treatment delivery, treatment planning, and other technical questions.

**Conclusions:** Incorporation of a medical physics patient consult program into clinical practice requires modest time commitment, and has the benefits of increasing medical physics engagement with patient care and improving patient satisfaction through better education.

## Introduction

Health literacy describes a patient’s ability to obtain and understand the information they need to make appropriate medical decisions. The Institute of Medicine has reported that approximately half of American adults have difficulty comprehending and utilizing health information^1^. Studies to understand this issue have shown that the process of acquiring health literacy starts with patients gaining proficiency in obtaining useful and accurate health information^2^. In the radiation oncology clinic, patients access health information primarily from the radiation oncologist with support from mid-level providers, nursing staff, and radiation therapists. This is typically supplemented by personal research on the internet and anecdotal advice from family and friends. Depending on the amount of time available for patient consults, providers may spend the majority of the time discussing topics directly related to the patient’s medical condition, leaving very little time to discuss other concerns^3^. During treatment, patients are confronted with sophisticated technology and complicated planning concepts that could lead to fear and anxiety. Clinical medical physicists are ideally placed to have a positive impact on health literacy by serving as an information resource and a guide through the complex treatment process.

Access to treatment related information has a strong impact on patient satisfaction scores in radiation oncology, and high patient satisfaction has positive influences on quality of care, including increased patient compliance, improved patient-physician relationships, and reduced anxiety^13,14^. Additionally, non-technical quality indicators in radiation oncology are incorporating patient satisfaction as a measure of quality from the patient’s perspective^15^. As a result, efforts to increase patient satisfaction will become increasingly significant for optimal quality of care.

The primary purpose of the present study was to develop and implement a new, formalized patient consult program in our clinic, where patients are invited to meet directly with a clinical medical physicist to learn and ask questions about the technical aspects of their care. Further, we aimed to evaluate the required time and effort of the program, as well as gain insight into the program benefits from a patient perspective.

## Methods

### A. Program development

In our practice, it was customary for clinical medical physicists to assist with patient education as needed. The typical opportunities for patient interaction included direct request from the radiation oncologist or other support staff, involvement with support group educational programs, or specific patient request. Until recently, these interactions occurred informally and were not incorporated into routine practice for our physics group. However, prompted by ongoing positive patient feedback and overall support from the clinical staff following our patient encounters, we decided to formalize our patient education initiatives into a medical physics patient consult. This concept was introduced to the department staff and various details pertaining to effective implementation were discussed, including consult placement into the clinical workflow, time commitment, and data collection.

### B. Data collection

The following data are collected following each patient encounter: age, gender, treatment intent, treatment site, consult duration, discussion points, overall impression, and a summary of the questions asked during the consult.

### C. Qualitative data analysis

The qualitative data analysis focused on understanding the number and types of questions asked during the patient consults. The collection of patient questions was sorted according frequency of occurrence and grouped into question types. The analysis also focused on evaluating the patient encounter notes for interesting insights regarding meeting tone, number of meeting attendees, and other non-clinical discussion topics.

## Results

### A. Consult placement in the clinic workflow

Patients are invited to meet with a clinical medical physicist on a voluntary basis directly after the treatment planning CT, and then again after treatment has started (Figure 1). The first meeting serves as a general education session, while the second meeting provides an opportunity for patients to review their treatment plans with a medical physicist. The rationale for doing this is 3-fold:

**Figure 1.**
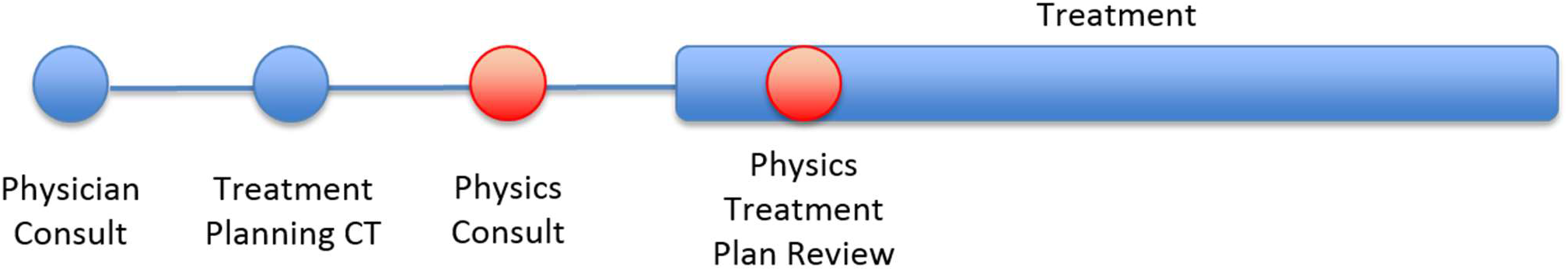
Placement of the medical physics consult and treatment plan review into the clinical workflow. Blue = existing standard patient care process, Red = placement of new medical physics interactions.

1. The patient has already had time to digest the information presented during the physician consult.
2. A medical procedure has occurred that might invite questions from patients.
3. Making the patient encounter voluntary allows the clinical team to become comfortable with the new process by not putting excessive burden on the clinical workflow.

### B. Patient consult details

Directly after the treatment planning CT, patients are invited to meet with a clinical medical physicist to review treatment planning and radiation delivery details. If the patient desires to have a meeting, they are placed in an exam room for private discussion. Family and friends are also invited to attend. Each new consult starts with explaining the purpose of the meeting as well as the clinical medical physicist’s role in patient care. We have also found it helpful to acknowledge with the patient that we know they are receiving a large volume of information about treatment, and that medical physicists are available to help them understand the technical aspects of their care. This sets a nice tone for the consult by helping patients feel comfortable and supported. The treatment planning CT process is then explained, followed by a detailed explanation of the treatment planning and radiation delivery processes. A general overview of what to expect in the treatment room is then reviewed. Patients are invited to ask questions at any point during the discussion. In general, the medical physicist will use patient cues to guide the level of detail appropriate for the discussion by assessing the level of patient engagement, question complexity, and prior knowledge of any background information about the patient, such as professional background. Any question outside of the scope of practice for a medical physicist is referred to the appropriate discipline for the answer. The general discussion details for the patient consults are summarized in Table 1.

**Table 1.**
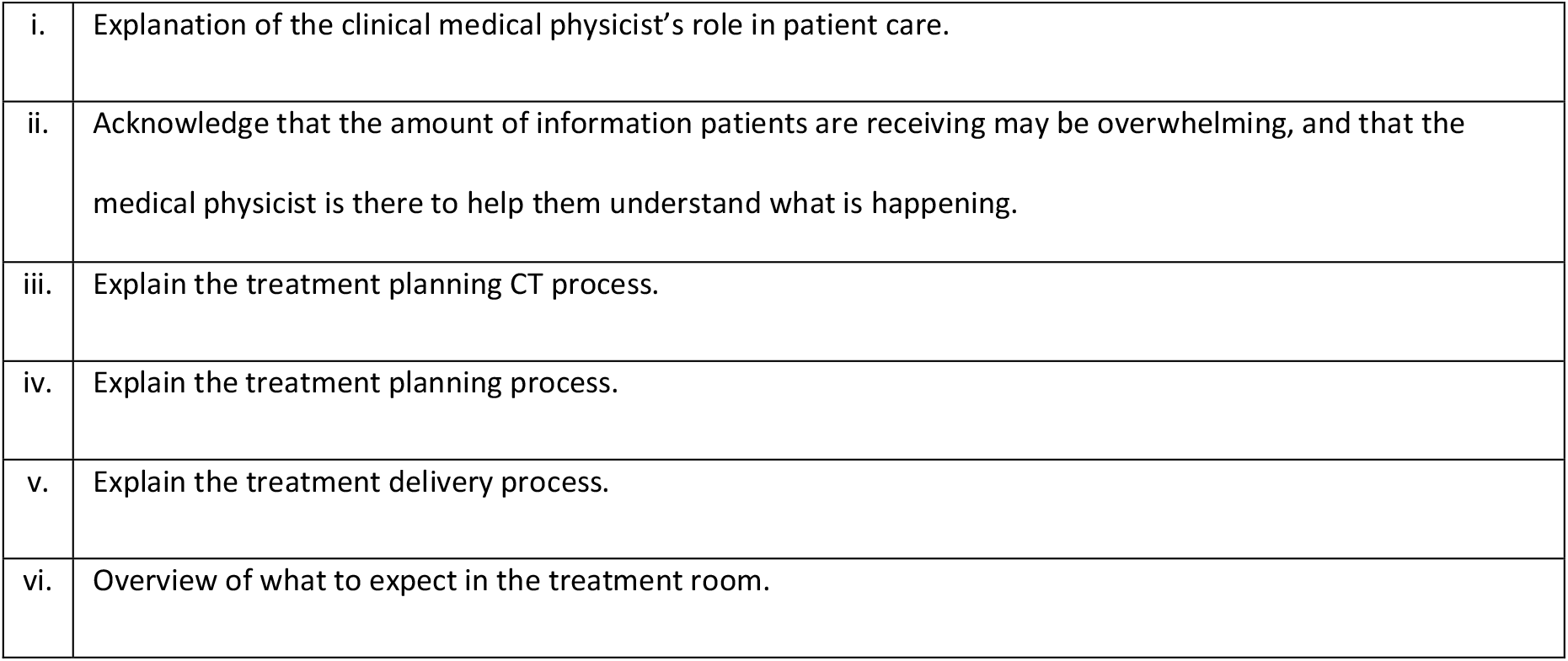
Typical discussion sequence during the medical physics patient consults.

|  |  |
| --- | --- |
| i. | Explanation of the clinical medical physicist's role in patient care. |
| ii. | Acknowledge that the amount of information patients are receiving may be overwhelming, and that the medical physicist is there to help them understand what is happening. |
| iii. | Explain the treatment planning CT process. |
| iv. | Explain the treatment planning process. |
| v. | Explain the treatment delivery process. |
| vi. | Overview of what to expect in the treatment room. |

### C. Summary of patient characteristics

A summary of patient characteristics is presented in Table 2.

**Table 2.** Summary of medical physics patient consult characteristics.

|  |  |
| --- | --- |
| Program Start | August 2016 |
| Number of Patient Consults (% of total department patient load) | 63 (29%) |
| # Male (% of total patient consults) | 29 (46%) |
| # Female (% of total patient consults) | 34 (54%) |
| **Disease Site** | **# (% of total patient consults)** |
| - Breast | 26 (41.3%) |
| - Prostate | 17 (27%) |
| - CNS | 7 (11.1%) |
| - GYN/GI | 6 (9.5%) |
| - Lung | 4 (6.3%) |
| - Head & Neck | 1 (1.6%) |
| - Skin | 1 (1.6%) |
| - I-131 | 1 (1.6%) |
| % Curative Intent | 88% |
| % Palliative Intent | 12% |
| Average Age | 66.2 y |
| Average Consult Length | 24 min |

### D. Summary of initial impressions

Part of the data collection effort for this project involved noting the overall impression of the meeting, taking specific note of overall tone, meeting attendees, and other discussion points such as personal stories or background. When evaluating the patient encounter notes for overall tone, 55 patients (87%) had an encounter note that used positive descriptors such as “pleasant conversation”. Many times, patients bring other people with them into the physics consults, and this happened for 33 patients (52%). A few of the more pleasant aspects of the patient consults are making personal connections and having non-clinical conversations. Twenty seven patients (43%) contributed personal stories or professional background information to the conversation. These conversation elements improved the overall tone of the consults, and in many cases, helped provide context for the level of technical detail that might be appropriate for the discussion. A summary of more general observations collected during the patient consults is presented in Table 3.

**Table 3.** Summary of general observations and initial impressions collected from the medical physics patient consults.

|  |  |
| --- | --- |
| i. | Patients welcome the opportunity to meet with a physicist. |
| ii. | The discussions are engaging and surprisingly technical. |
| iii. | Patients are thankful for the additional information. |
| iv. | Anxious patients seem to benefit a great deal from the consult. |
| v. | Medical physicists are more engaged with patient care due to increased visibility to the patients and the clinical staff. |
| vi. | The patient relationship extends beyond the consult room. Patients feel comfortable asking more questions if needed. |
| vii. | The medical physics patient consults have been a rewarding experience. |

### E. Analysis of patient questions

Sixty five distinct questions were collected during the patient consults. One hundred ninety seven total questions were asked, which include repeat questions from different patients. The top 15 most frequently asked questions are presented in Table 4. Further analysis was performed by grouping the data into question types as follows: treatment delivery, treatment planning, imaging, technical, quality & safety, post-treatment, education resources & information, side effects, and medical professional roles. The frequency of occurrence for each question type is presented in Figure 2 (% = number of questions in each type out of the total number of questions asked (197)). These data show that the majority of the consult time addresses questions related to treatment delivery, treatment planning, and other technical questions.

**Table 4.**
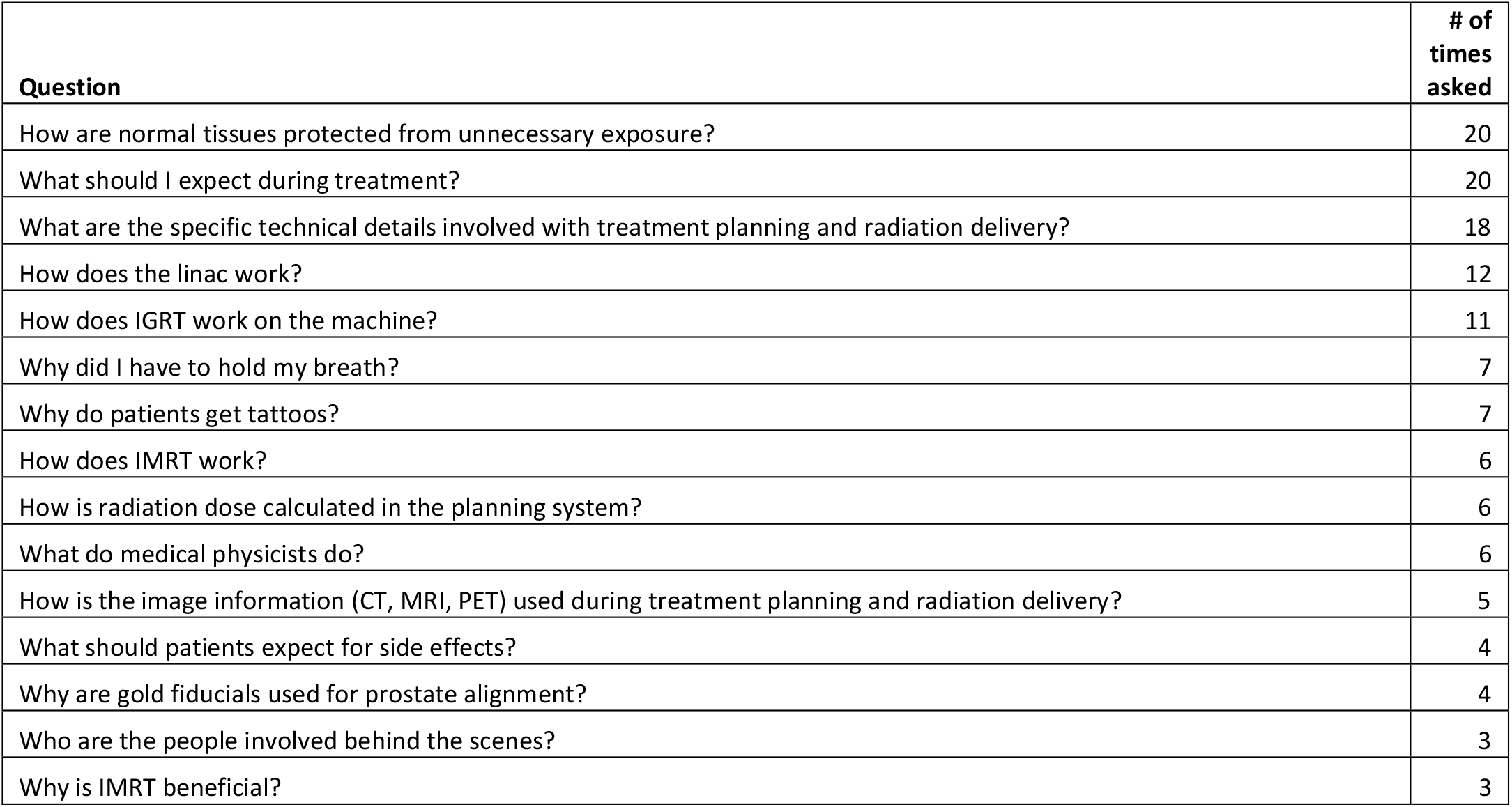
The 15 most frequently asked questions during the medical physics consults.

| Question | # of times asked |
| --- | --- |
| How are normal tissues protected from unnecessary exposure? | 20 |
| What should I expect during treatment? | 20 |
| What are the specific technical details involved with treatment planning and radiation delivery? | 18 |
| How does the linac work? | 12 |
| How does IGRT work on the machine? | 11 |
| Why did I have to hold my breath? | 7 |
| Why do patients get tattoos? | 7 |
| How does IMRT work? | 6 |
| How is radiation dose calculated in the planning system? | 6 |
| What do medical physicists do? | 6 |
| How is the image information (CT, MRI, PET) used during treatment planning and radiation delivery? | 5 |
| What should patients expect for side effects? | 4 |
| Why are gold fiducials used for prostate alignment? | 4 |
| Who are the people involved behind the scenes? | 3 |
| Why is IMRT beneficial? | 3 |

**Figure 2.**
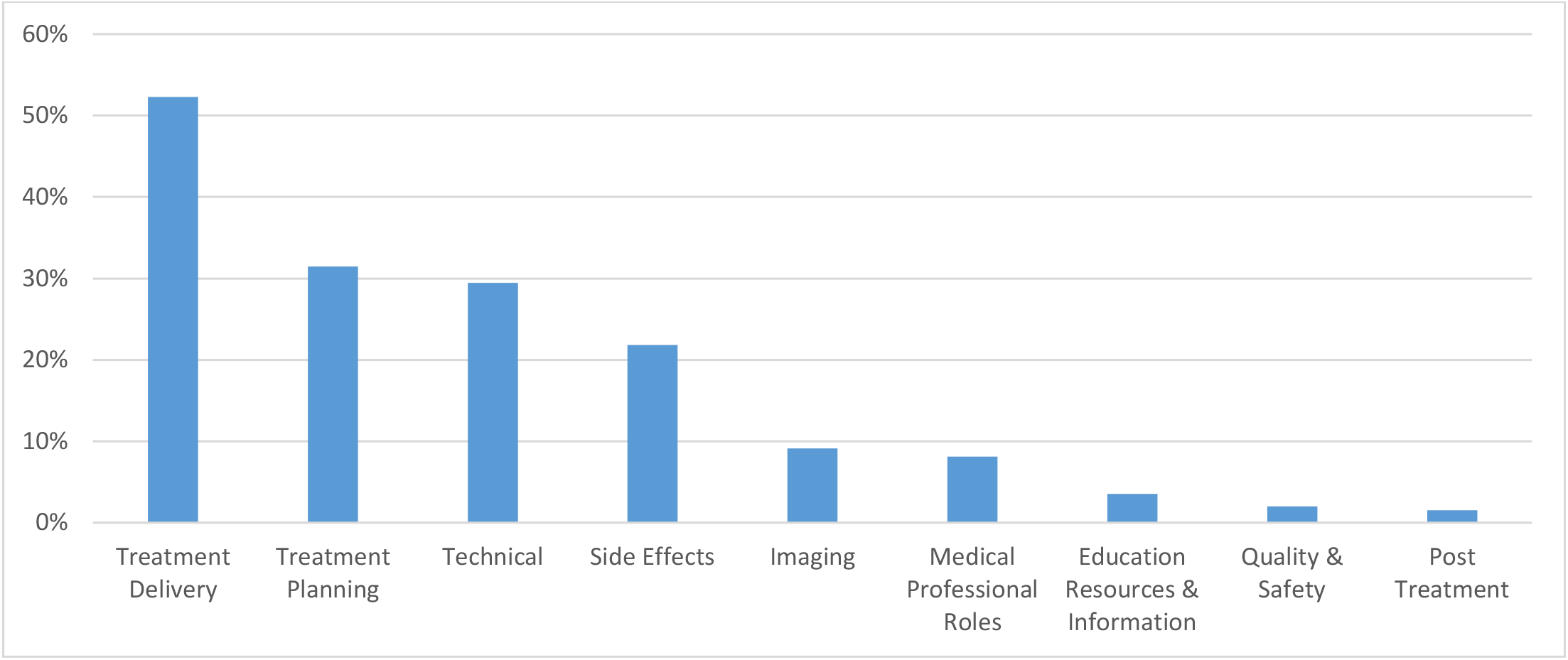
Frequency of occurrence for each question type during the medical physics consults. (% = number of questions in each type out of the total number of questions asked (197)).

## Discussion

This manuscript describes the development and implementation of a formalized medical physics patient consult program. This program represents an expansion of clinical medical physics practice into a new area of direct patient interaction^17^. Recent AAPM initiatives have called on medical physicists to explore new areas of clinical practice and public engagement by utilizing their technical expertise in new ways^10,16^. Atwood, et al. argue that direct patient care is a critical area for expansion that will establish new essential roles and responsibilities for clinical medical physicists^11^. The medical physics patient consult program described here is one of the first reports of community practice expansion made in the spirit of these recent initiatives.

This report presents a new opportunity for clinical medical physicists to improve patient care by helping to establish trust, provide information, and reduce anxiety. Oncology patients face a diverse, multi-disciplinary landscape during treatment. They encounter a wide array of diagnostic tests, specialty consults, reports, and in some cases, alternative care options. This, coupled with many patients’ limited understanding of the basic concepts related to cancer therapy, may evoke anxiety and feelings of being overwhelmed^4^. Such elevated emotional states may impair the patient’s learning process by creating an intellectual block to comprehending the information being presented^5^. As patients seek information about their care, they balance limited access to provider-based information with an abundance of online information sources, many of which could be inaccurate^5^ or too complex^6,7,8^. Increased awareness of these difficulties has led to improved patient support programs and attempts to better understand the impacts of patient distress and health literacy on outcomes^9,2^. Widespread adoption of clinical medical physics consults into clinical practice may help reduce patient anxiety and improve the patient’s learning process, potentially contributing to increased satisfaction with their care^17^.

The need for more information is a significant predictor for decreased patient satisfaction^13,14^. Geinitz, et al. showed that within the first week of radiation therapy, 46.1% of patients wished they had received additional information before the onset of treatment^13^. This provides reinforcement for the timing and placement of our physics consult directly after the treatment planning CT before treatment starts. Furthermore, the Geinitz study reported that the most requested information items pertained to radiation delivery and x-ray effects: “How do x-rays carry out their effect?” (65.2% of requests), “Function of the linear accelerator.” (59.8%), “What are x-rays?” (52.1%), and “Side effects.” (45.3%)^13^. The majority of the requested additional information is technical in nature and matches the overall trends in our patient questions data. Interestingly, both of our datasets show that side effects remain a primary concern for patients even after the initial physician consult.

The most notable impression of this new program is that patients welcome the opportunity to meet with a clinical medical physicist. Overall, the discussions are engaging and surprisingly technical. Discussion topics have ranged from detailed reviews of linear accelerator operations with patients who are engineers, all the way to techniques for reducing normal tissue exposure for nervous breast cancer patients. In general, anxious patients seem to benefit a great deal from consultation with a clinical medical physicist. We believe that the patient’s ability to have a dedicated discussion about the technical aspects of their care, especially pertaining to radiation delivery, provides a meaningful benefit to patients who are nervous about what to expect in the treatment room. Unsurprisingly, these patients express gratitude for the additional information.

The array of patient questions in our study is surprisingly diverse. This may indicate that patients are undergoing a process of understanding the medical information being presented according to recent models of health literacy, which describe how patients obtain and understand new medical information^2^. Sixty five distinct questions were asked from a study cohort of 63 patients. Even though the majority of the discussion points fit within the scope of practice for a clinical medical physicist (Figure 2), it is clear that patients are also seeking answers to questions that were likely already addressed during the physician consult or discussions with other providers (e.g., questions regarding treatment planning or treatment side effects). This type of repeated inquiry to the department staff fits into the model of health literacy described by Sorensen, et al., which explains that within the domain of healthcare, patients achieve health literacy by accessing, understanding, processing, and applying information relevant to their care^2^. Therefore, to support this learning process, patients are taking advantage of the medical physics consult to also ask a broad set of questions about their care, in addition to the focused medical physics questions seen in our data. Furthermore, spaced repetition of education information improves the efficiency and effectiveness of learning, and a consult with a clinical medical physicist contributes to the patient’s learning process by not only providing a new source of information but also additional access to repeated information over a spaced time interval^12^.

The increased visibility of the clinical medical physicist to the patient has changed the way medical physicists are perceived in general around the department. The patient relationship extends beyond the consult room. Patients immediately recognize the medical physicist as an integral part of their treatment team, and they feel comfortable asking more questions, either through casual conversations or additional scheduled meetings. This program has also enriched the relationship between the physicist and the rest of the clinical staff. The nursing and radiation therapy staff routinely engage the medical physicist with direct patient care matters, usually pertaining to further patient questions about treatment delivery after treatment starts. The professional relationship between the radiation oncologists and medical physicist has evolved in a positive way by knowing that a medical physicist is available for patient consultation if needed. The medical physicists now get direct referrals from the radiation oncologists to meet with the patients they feel will benefit from a more technical discussion. Overall, the medical physics staff is more engaged with patient care, and this has resulted in greater visibility to the entire clinical staff and management team.

Our patient consult program was started without consideration for medical physics training needs and billing matters. Clinical medical physicists do not routinely receive professional training on how to engage and interact with patients. A good place to gain experience meeting with patients is to shadow the radiation oncologists during their initial consults or weekly on-treatment visits. Another place to gain experience talking about technical concepts in front of patients is to deliver educational talks to patient support groups. We found this to be very beneficial in helping medical physicists gain experience translating complex technical concepts into simpler language for patients. In general, it is likely that training needs will need to be addressed at the medical physics residency level if direct patient consultation is to be incorporated into formal medical physics practice on a large scale^18^. Further, concerning the financial implications, the clinical medical physics time with the patient was not billed. In an era of increasing efficiency in healthcare delivery, this non-billable clinical time could be a complicating factor to implementation for some busy or resource-limited clinics.

In conclusion, incorporation of a formalized medical physics patient consult program into clinical practice requires modest time commitment, and has the benefits of increasing medical physics engagement with patient care and improving patient education, potentially leading to increased patient satisfaction.

## Data Availability

Data available from the corresponding author.

## References

1. Institute of Medicine. Health Literacy: a prescription to end confusion. Washington DC: The National Academies; 2004.

2. Sorensen K, Van den Brouche S, Fullam J, et al. Health literacy and public health: A systematic review and integration of definitions and models. BMC Public Health. 2012; 12:80.

3. Siminoff LA, Graham GC, Gordon NH. Cancer communication patterns and the influence of patient characteristics: Disparities in information-giving and affective behaviors. Patient Education and Couseling. 2006; 62, 255–360.

4. Matsuyama RK, Lyckholm LJ, Molisani A, Moghanaki D. The value of an educational video before consultation with a radiation oncologist. J Cancer Educ. 2013; 28(2), 306–313.

5. Dauer LT, Thornton RH, Hay JL, Balter R, Williamson MJ, St. Germain J. Fears, Feelings, and Facts: Interactively Communicating Benefits and Risks of Medical Radiation with Patients. Am J Roentgenol. April 2011; 196(4).

6. Rosenberg SA, Francis DM, Hullet CR, et al. Online Patient Information from Radiation Oncology Departments is Too Complex for the General Population. Practical Radiation Oncology. 2017; 7, 57–62.

7. Prabhu AV, Hansberry DR, Agarwal N, Clump DA, Heron DE. Radiation Oncology and Online Patient Education Materials: Deviating from NIH and AMA Recommendations. Int J Radiation Oncol Biol Phys. 2016; 96(3), 521–528.

8. Prabhu AV, Crihalmeanu T, Hansberry DR, et al. Online palliative care and oncology patient education resources through Google: Do they meet national health literacy recommendations? Practical Radiation Oncology. 2017; 7, 306–310.

9. Habboush Y, Shannon RP, Niazi SK, et al. Patient-reported distress and survival among patients receiving definitive radiation therapy. Advances in Radiation Oncology. 2017; 2, 211–219.

10. AAPM Med Phys 3.0; https://www.aapm.org/MedPhys30/

11. Atwood TF, Brown DW, Murphy JD, Moore KL, Mundt AJ, Pawlicki T. Care for Patients, Not for Charts: A Future for Clinical Medical Physics. Int J Radiation Oncol Biol Phys. 2018; 100(1), 21–22.

12. Kang SHK. Spaced repetition promotes efficient and effective learning: Policy implications for instruction. Policy Insights from the Behavioral and Brain Sciences. 2017; 3(1), 12–19.

13. Geinitz H, Marten-Mittag B, Schafer C, et al. Patient satisfaction during radiation therapy. Strahlenther Onkol. 2012; 188, 492–498.

14. Lam WWT, Kwong A, Suen D, et al. Factors predicting patient satisfaction in women with advanced breast cancer: a prospective study. BMC Cancer. 2018; 18, 162.

15. Albert JM, Das P. Quality indicators in radiation oncology. Int J Radiation Oncol Biol Phys. 2013; 85(4), 904–911.

16. AAPM Public Education; https://www.aapm.org/org/structure/default.asp?committee_code=PE

17. Atwood TF, Brown DW, Murphy JD, Moore KL, Mundt AJ, Pawlicki T. Establishing a new clinical role for medical physicists: A prospective phase II trial. Int J Radiation Oncol Biol Phys. 2018; 102(3), 635–641.

18. Brown DW, Atwood TF, Moore KL, MacAulay R, Murphy JD, Mundt AJ, Pawlicki T. A program to train medical physicists for direct patient care responsibilities. J Appl Clin Med Phys. 2018; 19(6), 332–335.

